# Identification of targets for drug repurposing to treat COVID-19 using a Deep Learning Neural Network

**DOI:** 10.1101/2023.05.23.23290403

**Authors:** Si-Han Wang, Yu-Hsuan Tang, Hao Hsu, Chu-Nien Yu, Oscar Kuang-Sheng Leea

## Abstract

The COVID-19 pandemic has resulted in a global public health crisis requiring immediate acute therapeutic solutions. To address this challenge, we developed a useful tool deep learning model using the graph-embedding convolution network (GECN) algorithm. Our approach identified COVID-19-related genes and potential druggable targets, including tyrosine kinase ABL1/2, pro-inflammatory cytokine CSF2, and pro-fibrotic cytokines IL-4 and IL-13. These target genes are implicated in critical processes related to COVID-19 pathogenesis, including endosomal membrane fusion, cytokine storm, and tissue fibrosis. Our analysis revealed that ABL kinase inhibitors, lenzilumab (anti-CSF2), and dupilumab (anti-IL4Rα) represent promising therapeutic solutions that can effectively block virus-host membrane fusion or attenuate hyperinflammation in COVID-19 patients. Compared to the traditional drug screening process, our GECN algorithm enables rapid analysis of disease-related human protein interaction networks and prediction of candidate drug targets from a large-scale knowledge graph in a cost-effective and efficient manner. Overall, Overall, our results suggest that the model has the potential to facilitate drug repurposing and aid in the fight against COVID-19.

## 1. Introduction

Coronavirus disease 2019 (COVID-19) emerged in December 2019 and has rapidly spread worldwide, resulting in a global public health crisis. Severe acute respiratory syndrome coronavirus 2 (SARS-CoV-2), which causes COVID-19, is a single-stranded positive RNA virus with spikes on its envelope [1]. As of 5 April 2023, the World Health Organization COVID-19 Dashboard has reported 762,201,169 confirmed cases and 6,889,743 deaths worldwide. Many patients develop pneumonia within weeks of showing symptoms and increased inflammatory cytokines which can lead to respiratory failure and death [2]. The traditional drug discovery method, with a long discovery period (10–15 years) and low success rate (2.01%) [3], has failed to meet the urgent need for a COVID-19 cure.

Numerous monoclonal antibodies are currently being developed as systemic therapies for COVID-19; however, their large size, instability, and low density of binding sites (two per 150 KDa antibody) make them unsuitable for intranasal delivery [4]. To overcome this challenge, deep learning, a subset of artificial intelligence (AI) algorithms, can accelerate drug development and repurposing to slow down the progression of acute disease. Deep learning uses multiple interconnected layers to recognize complex patterns in data, which is useful for finding correlations between inputs and outputs for complex problems. Unlike the traditional protein structure prediction problem, locally connected graph neural networks can accurately model the structure-to-sequence mapping problem [5]. The Graph Convolutional Network (GCN) is an approach to graph embedding that transforms graph information into spectral domains for convolutional calculation. This is done using the Laplacian matrix, Chebyshev polynomials, or other methods [6-10]. GCNs are powerful in processing network data, and their representative tasks include node classification, link prediction, and graph generation., as they can combine features with a hidden layer to aggregate important information from different nodes. By combining features with a hidden layer, GCNs can effectively aggregate important information from different nodes. This ability to recognize complex patterns in data and find correlations between inputs and outputs for complex problems makes GCNs an excellent choice for prediction models. Additionally, GCNs’ locally connected graph neural networks can accurately model the structure-to-sequence mapping problem, providing greater accuracy compared to traditional protein structure prediction methods. Moreover, recent studies have shown that deep learning, including GCNs, outperforms classic machine learning methods in assisting drug repurposing, allowing the screening of existing drugs as potential treatments for SARS-CoV-2. The development of affordable approaches for the effective treatment of COVID-19 is challenging without foreknowledge of the complex networks connecting drugs, targets, SARS-CoV-2, and diseases. Current studies suggest that using the in-silico method to identified some potential antiviral drugs such as remdesivir [11, 12], mefuparib [13], and toremifene [14, 15], while drugs like baricitinib [16], melatonin [17], and dexamethasone [18, 19] have been identified as potential host-targeted therapies to attenuate cytokine storms in COVID-19 patients. The results of clinical trials for these drugs are controversial [20]. However, most of these studies focused on SARS-CoV-2 proteins with known functions, such as the spike protein and 3C-like (3CL) protease, or a host protein with a known interaction with viruses, such as ACE2 and TMPRSS2. The approach of focusing only on SARS-CoV-2 proteins and host proteins with known interactions with viruses significantly limits the potential targets for drug development because the role and functional annotations of many other host proteins are mostly ignored. In this study, we propose a new AI-assisted drug development method, the graph-embedding convolution network (GECN), that utilizes the functional annotations of host proteins to identify potential druggable targets for SARS-CoV-2 pathogenic mechanisms. By investigating tissue specificity, physiological functions, and cell signaling pathways, our approach aims to expedite drug discovery new insights into the druggable targets and pathogenic mechanisms of COVID-19.

## 2. Materials and Methods

### 2.1 Data preprocessing

The protein interaction data were extracted from the STRING dataset (v 11.0) [21] with ***Homo sapiens*** axotomy (ID 9606), and edges between pairs of all associated genes were constructed, regardless of whether it was physically binding or not. The edges were not weighted, and no edge attributes were added. GO terms, such as cell components, biological processes, and molecular functions, were extracted and clustered using the semantic similarity method [22, 23]. In our study, semantic similarity was represented by the minimum number of steps across the graph required to connect two terms, weighted by how specific or general the terms are, and was calculated with the following formula (2):

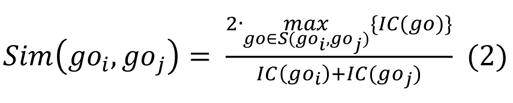

where go_i_ and go_*j*_ are a pair of GO terms and Sim(go_i_, go_j_) are their semantic similarities. S(go_i_, go_j_) is the subset of GO terms shared between go_i_ and go_*j*_ after propagating up the GO DAG using the “is_a” and “part_of” relations. *IC*(go_*j*_) is the information content (IC) of a GO term, which is defined as the frequency of go_*j*_, relative to the total number of GO terms in the UniProt Gene Ontology Annotation (GOA) database [24, 25], specifically calculated using the formula (3):

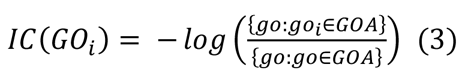

Pathway IDs from Reactome were clustered by propagating along the Reactome [26, 27] graph and were mapped to the top super pathway. Finally, the “probability of loss of function intolerance” extracted from ExAC (v 1.0) [28] was normalized.

### 2.2 Graph convolution network (GCN)

The GCN algorithm included a feature matrix and an adjacency matrix [9]. A feature matrix was N × F⁰ dimension, with N being the number of nodes and F⁰ being the feature numbers in each node. An adjacency matrix represented the structure of the graph, and the dimension was N x N.

Each neural network layer can be written as a non-linear function (4)[29], as shown below, where H (0) = X and H(l) = Z (or z for graph-level outputs), and l is the number of layers.

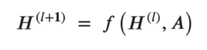

Here, W(l) is a weight matrix for the l-th neural network layer. Additionally, Â = A + I, where I is the identity matrix, *D*^ is the diagonal node degree matrix of Â, and σ(·) is a non-linear activation function, like ReLU.

Besides, we use multilayer GCN to combine neighboring node features with self-loops by summing and taking weighted average of their feature vectors which employ a multilayer GCN with the following layer-wise propagation rule:

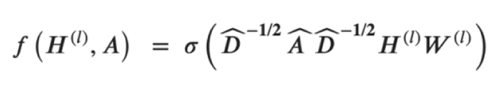

The Variational Graph Autoencoder (VGA) is a type of neural network used for generating molecular graphs and consists of an encoder and a decoder. The encoder of the VGA, which employs edge condition convolution, embeds the original graph into a continuous vector space. The decoder generates a probabilistic fully connected graph according to the predefined number of nodes and updates the parameters through approximate graph matching to improve the reconstruction ability of the autoencoder. We constructed the VGA model using three layers of Graph Convolutional Network (GCN) encoder with hidden unit dimensions of 512, 256, and 256, respectively. We used an inner product decoder to produce the output. The VGA is a type of Variational Autoencoder (VAE) that projects input features into a multivariate latent distribution and samples the values of latent features from this distribution. To make the sampling process differentiable and enable reliable training of the model, the reparameterization trick is used. The reparameterization trick involves sampling from a standard normal distribution and then transforming the sampled values to produce the latent features. The formula for the reparameterization trick is given as (1):

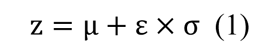

where ε∼N(0,1) and z is the latent feature. By using the reparameterization trick, the model can backpropagate gradients through the sampling process, allowing for end-to-end training of the variational autoencoder.

A neighbor sampler with a batch size of 1,024 was applied, and the loss function contained reconstruction loss and KL loss. Reconstruction loss is the cross-entropy loss of positive edges (interactions) and negative edges (non-interactions), whereas KL loss is the KL divergence between the output distribution of encoder N(μ, σ) and standard normal distribution *N*(0,1). The AdamW optimizer was used [30], and 100 training epochs were trained without early termination. Finally, the model with the best AUC value was saved.

To predict potential links and appropriate evaluation of the performance of our model, fixed-threshold metrics and test set sampling were required. We sampled those negative edges where the geodesic distances were equal to 2. The shortest paths between these pairs of nodes were equal to two, and improvements in these pairs were weighted more than at greater distances. To decrease the computational complexity of spectral graph convolution, the Chebyshev polynomial was used to replace the eigen-decomposition of a Laplacian matrix [31]. The Chebyshev polynomial utilized the recurrence relation to approach the value of eigen-decomposition and increase the efficiency of model training. The hyperparameter k in the Chebyshev convolution represents a receptive field that can lead the algorithm to see more local values. In our model, we set k to 2. Cluster-GCN is a type of GCN algorithm that is suitable for training by exploiting the graph clustering structure [32], In this study, a graph clustering algorithm identified a block of nodes associated with a dense subgraph. It then restricted the neighborhood search within this subgraph. This simple but effective strategy led to significantly improved memory and computational efficiency while achieving a test accuracy comparable to that of previous algorithms.

### 2.3 K-fold cross-validation

K fold cross-validation was used to verify the performance of the model. It split the data into five groups and randomly selected one of them as the test set. This approach estimated the performance of the model using the new data.

### 2.4 Protein-protein docking using MOE software

*In silico* protein-protein docking of ABL1 against TNF and ABL2 against BTK was performed using the MOE software [33]. PDB files of ABL1 (PDB ID: 1AWO), TNF (PDB ID: 1TNF), ABL2 (PDB ID: 5NP3), and BTK (PDB ID: 5XYZ) were downloaded from the RCSB PDB and imported into the MOE software. The pre-bound ligands were first removed from the PDB structures. Protein structures were then prepared using the MOE “Protein Preparation” tool (to correct atom lost issues) and MOE “Protonate 3D” tool (to add hydrogen atoms and to assign ionization states throughout the system). Water molecules farther than 4.5 Å from proteins were deleted. Active sites of TNF and BTK were found using the MOE “Site Finder” tool (Compute, Site Finder), and ABL1 or ABL2 were docked to all these sites later. Prepared structures were docked by MOE “Dock” tool (Compute, Dock, Protein-protein). Docking structures with a minimum S score indicated that the highest binding affinity was selected for further interaction analyses. Interaction bond type, bond length, and bond energy were indicated by MOE “Protein Contacts” tool (Protein, Protein Contacts).

## 3. Results

### 3.1 Experiment setting of the graph embedding convolution network algorithm

The GECN algorithm consists of two deep learning methods: GE and GCN, as shown in Fig. 1. The GE model uses a Variational Graph Autoencoder (VGA) to extract latent features from raw data, which are then used as input node features for the second deep learning model, DeepGCN, to perform GCN as a classifier. The VGA encoder maps raw data into a latent distribution with means and standard deviations, and a reparameterization trick is employed to make the VGA suitable for backpropagation. This makes the process of stochastic sampling of latent features from the latent distribution differentiable. The decoder of the VGA reconstructs the latent features into the adjacency matrix. The unsupervisedly trained VGA utilizes the reconstruction error and Kullback-Leibler divergence to encode latent features, which are then fed as node feature inputs into the supervisedly trained DeepGCN for classification. In brief, DeepGCN utilizes a combination of latent features as nodes, PPI network as graph structure, and disease-related genes as labels for training. The model is then trained with focal loss to minimize the difference between predicted and true labels and is enhanced with residual and dense connections for improved performance in comparison to traditional GCN with a deeper structure. It predicts potential protein targets that interact with COVID-19.

**Fig. 1.**
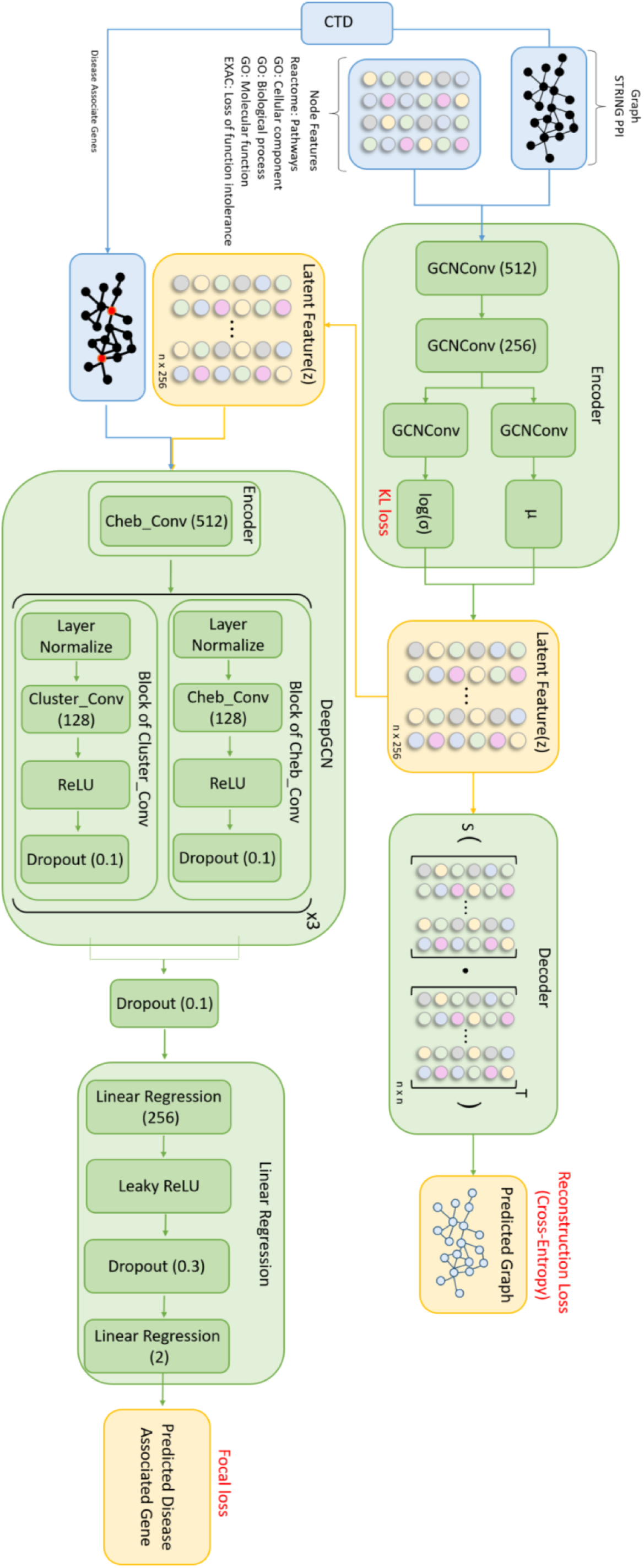
Overview of the graph-embedding convolution network (GECN) algorithm. The GECN algorithm combines graph embedding and graph convolution network. A variational graph autoencoder learned embedding gene features and protein interaction networks to latent features. DeepGCN predicts disease-associated genes based on latent features.

In this study, we conducted experiments on a biological network dataset consisting of a total of 533,138 data points. These data points were divided into a training set and a test set. The training set contained 479,825 positive edges, representing interactions between pairs of genes. The test set included 53,313 positive edges and an equal number of negative edges, representing the absence of interactions between pairs of genes. To select the negative edges, we chose unconnected node pairs that had a geodesic distance of two. This sampling strategy was employed to mitigate the effects of class imbalance and to allow for a more accurate assessment of the model’s performance in link prediction.

We employed a VGA to compress the data and retrieve the latent features. The VGA was evaluated on a complete version of the graph without removing any edges. To train the model, we used the Adam optimization algorithm with an initial learning rate set to a predefined value. The batch size was fixed at 1024, and we trained the model for a total of 100 epochs. To mitigate the risk of overfitting, we applied L2 regularization (weight decay) with a specified weight value. The learning rate adjustment function used was the cosine annealing function, and the loss function employed was the cross-entropy loss function. During training, we calculated the loss between the predicted values and the ground truth. The model’s parameters were then optimized by the Adam optimization algorithm based on the calculated loss. To address the data imbalance problem, we used a cost matrix to give more attention to false positives and omissions.

### 3.2 Performance of the graph embedding convolution network algorithm

The performance of our deep learning model was evaluated using various metrics, including the area under the receiver operating characteristic curve (AUC) and fixed-threshold metrics such as accuracy, recall, precision, F1 score, and F2 score. The test set achieved an AUC of 0.92 (Fig. 2a), while the accuracy of the model with the threshold applied was 0.84, with a recall of 0.87 and a precision of 0.82 (Fig. 2b). The F1 and F2 scores were 0.84 and 0.86, respectively. The confusion matrix of the test set revealed 10,488 new predicted links, suggesting potential interactions between pairs of genes that were not constrained to physical binding due to the nature of the STRING dataset and may include metabolic pathways or cellular processes.

**Fig. 2.**
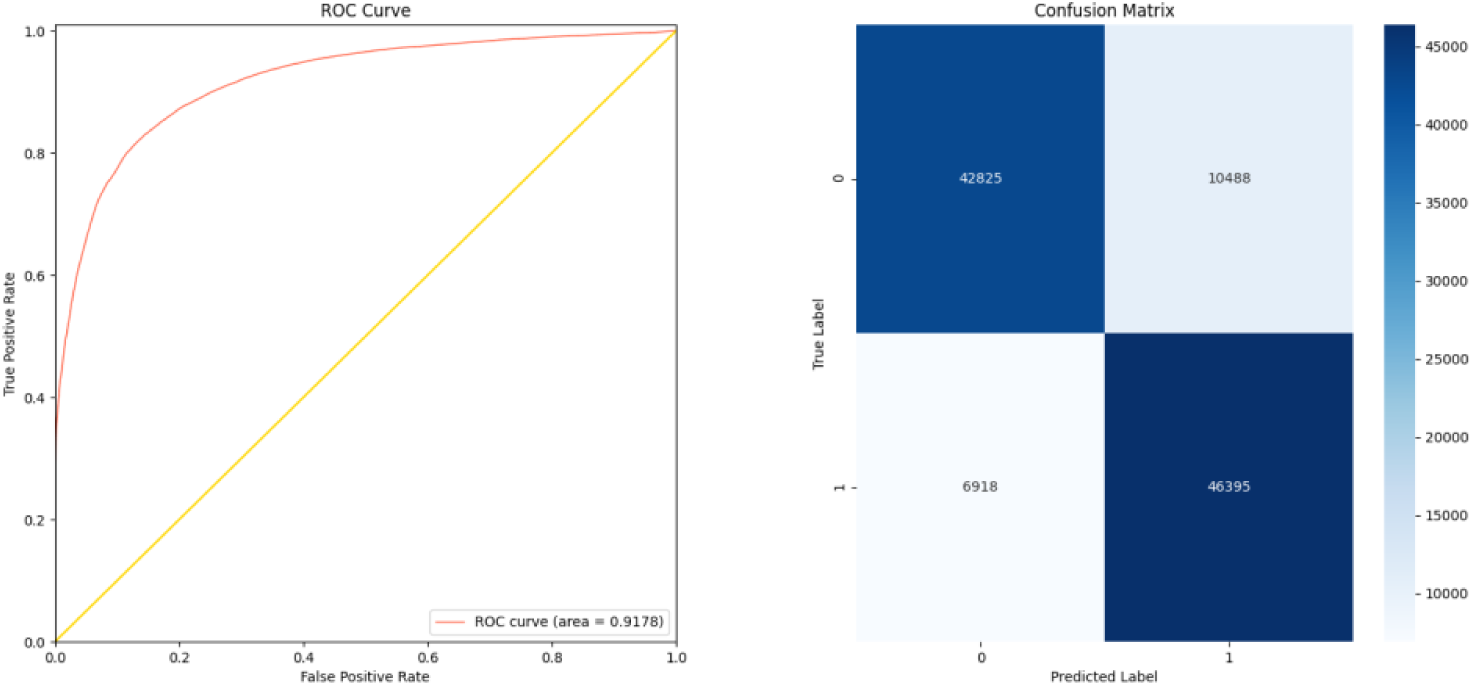
Evaluation of link prediction of the protein interaction network. The receiver operating characteristic (ROC) curve (**a**) and confusion matrix. (**b**) of link prediction were evaluated from the test set.

### 3.3 Predicting and Assessing Protein-Protein Interactions between COVID-19 and Associated Genes through Molecular Docking Analysis

To understand virus-host protein interactions have found that SARS-CoV-2 can interfere with various signaling pathways in host cells. The immune evasion of SARS-CoV-2 improves viral survival and triggers pathogenic mechanisms in host cells. Analyzing the host protein associated with the known COVID-19 related genes can reveal new druggable targets and can help heal COVID-19 patients by blocking pathogenic signaling pathways or potential virus-host protein interactions. Our analysis identified several predicted interacting genes that showed potential associations with COVID-19-related genes ABL1 (39.13%) and ABL2 (26.09%). Less than 1% of genes interacted with other COVID-19-related genes (see Supplementary Table 1). Notably, ABL1-TNF and ABL2-BTK had the highest probabilities of predicted links for ABL1 and ABL2, respectively.

To estimate the likelihood of physical binding in the predicted association, we used the Molecular Operating Environment (MOE) software [34, 35]. We selected the links with the highest probabilities of prediction and the docking structures of ABL1-TNF and ABL2-BTK as candidates. Our analysis revealed that the best docking structure of ABL1-TNF achieved an S score of -67.63 and a root-mean-square deviation (RMSD) of 0.98, while ABL2-BTK achieved an S score of -51.39 and RMSD of 1.16, indicating a high probability of physical interaction between these two pairs.

The 2D interaction diagram (Fig. 3) shows the three phenomena. First, amino acid residues are exposed in solution. Second, binding pockets are formed. Finally, the important amino acid residues are involved in protein interactions. For example, the side chain of Arg131 on TNF forms a hydrogen bond with the side chain of His95 on ABL1 (bond length = 3.345 Å, energy = 4.500 kcal/mol), the backbone of Gln88 on TNF forms a hydrogen bond with the side chain of Asn94 on ABL1 (bond length = 3.205 Å, energy = 3.100 kcal/mol), and the side chain of Glu23 on TNF forms an ionic bond with the side chain of Lys87 on ABL1 (bond length = 4.006 Å, energy = 0.501 kcal/mol). The backbone of Glu640 on BTK forms a hydrogen bond with the side chain of Lys87 on ABL2 (bond length = 3.439 Å, bond energy = 1.300 kcal/mol), and the side chain of Gln496 on BTK forms a hydrogen bond with the side chain of Arg103 on ABL2 (bond length = 3.199 Å, bond energy = 0.700 kcal/mol).

**Fig. 3.**
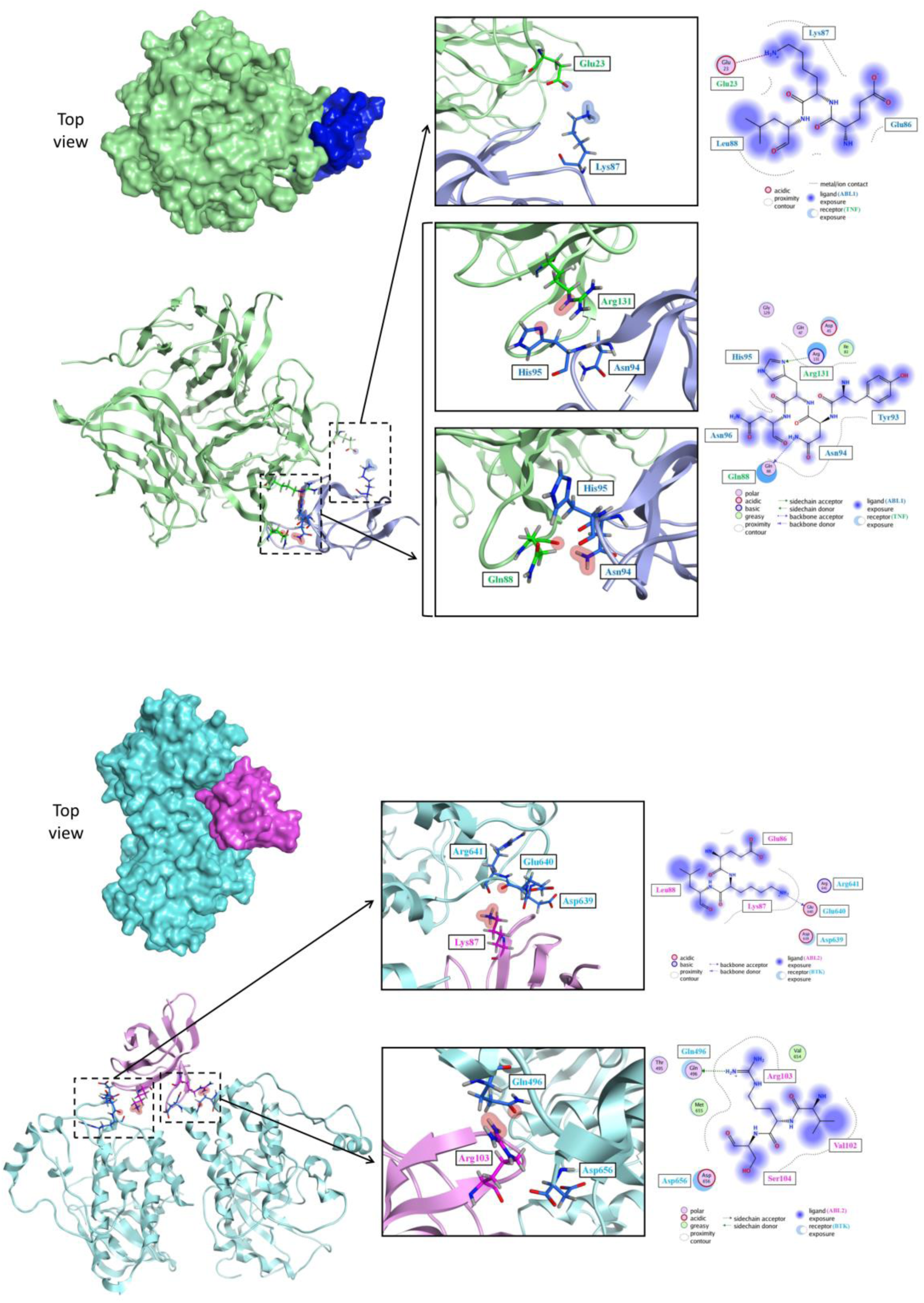
Docking structure with the lowest energy simulated by Molecular Operating Environment. The docking positions of the two proteins are clearly shown in Fig. 3 in the protein surface diagram (upper left panel) and ribbon diagram (upper right panel). **(a)** Docking structures of ABL1-tumor necrosis factor (TNF) and ABL1 (Protein Data Bank (PDB) ID: 1AWO) are shown as blue ribbons, whereas those of TNF (PDB ID: 1TNF) are shown as green ribbons; the skeletons are binding sites. (**b)** Docking structures of ABL2-Bruton’s tyrosine kinase (BTK) and ABL2 (PDB ID: 5NP3) are shown as pink ribbons and those of BTK (PDB ID: 5XYZ) are shown as green ribbons; the skeletons are binding sites.

### 3.4 Predicting Potential Target Genes in SARS-CoV-2 Affected Host Protein Networks using GECN Algorithm

The GECN algorithm was tested using 15 COVID-19 associated genes from CTD in 2019 as a graph center to construct SARS-CoV-2 affected host protein networks. The algorithm embedded the latency features from the autoencoder, with an accuracy of 0.99 ± 0.01 (mean ± SD), precision of 0.64 ± 0.11, recall of 0.77 ± 0.15, and F1 of 0.69 ± 0.08. The top five predicted targets were Interleukin (IL)-4, Colony-stimulating factor 2 (CSF2), IL-13, CXCL8, and PRL; CXCL8 was confirmed as a COVID-19 associated gene in 2021.

In the second stage, COVID-19 associated genes from CTD in 2021 were used as the graph center to construct a SARS-CoV-2 affected host protein network. The algorithm embedded the latency features from the autoencoder, with an accuracy of 0.99 ± 0.01 (mean ± SD), precision of 0.69 ± 0.06, recall of 0.80 ± 0.10, and F1 of 0.74 ± 0.06. The top three predicted targets were CSF2, IL-4, and IL-13. These target genes include ALB1/ALB2 (which might be involved in the entry of SARS-CoV-2 into host cells), CSF2 (which might participate in the cytokine storm, inducing severe syndrome), and IL-4 and IL-13 (which might participate in pulmonary fibrosis).

## 4. Discussion

The efficacy and safety of the COVID-19 pandemic has resulted in significant societal challenges. However, clinicians have no drug recommendations without a proper understanding of the virus and its mechanisms. In this context, in silico approaches led to the advancement of machine learning techniques and docking methods in drug repurposing. In this study, we developed a deep learning algorithm, the GECN, to predict COVID-19 related genes integrated with functional annotations of human genes. However, focusing on specific targets such as spike protein, 3CL protease, and MPro in SARS-CoV-2 or ACE2 and TMPRSS2 would restrict the candidate screening scope. Most interactive proteins in humans are associated with endomembrane compartments or vesicle trafficking pathways, indicating the essential role of human proteins during infection. Therefore, this study identified 332 physical interactions which provide form COVID-19 Research Group of Quantitative Biosciences Institute between human proteins and SARS-CoV-2 proteins, providing a comprehensive view of the host-pathogen interactome.

Our GECN algorithm predicted ABL1, ABL2, CSF2, IL-4, and IL-13 as candidates for COVID-19 treatment target genes, which were successfully identified as having a binding affinity with COVID-19-related proteins. The use of deep learning algorithms, such as the GECN used in this study, can be a powerful tool for analyzing large amounts of data and identifying patterns that may not be apparent to humans. Previous studies [36, 37] on the ABL family and coronavirus the ABL kinase inhibitor, GNF2, GNF5 [36], imatinib [36, 37], imatinib mesylate [38], nilotinib [38], and dasatinib [38] can reduce the viral titer of coronavirus by preventing syncytia formation. These results also suggest that ABL1 and ABL2 are involved in the process of coronavirus infection. Experiments on ABL kinase inhibitor treatment of SARS-CoV-2 have shown promising results that imatinib mesylate suppresses the replication of SARS-CoV-2 in Vero-E6 cell[39]. Moreover, study on nilotinib, dasatinib, and imatinib reported antiviral activity only of nilotinib but not of dasatinib or imatinib in Vero-E6 cells and Calu-3 cells [40]. Imatinib did not exhibit any antiviral activity in Caco-2 cells, as documented in a previous study [41]. Three randomized clinical trials are currently underway to study imatinib’s efficacy in treating COVID-19. NCT04357613 (France), NCT04394416 (USA), and EudraCT2020-001236-10 (The Netherlands), are currently underway to study the therapeutic efficacy of imatinib in COVID-19 patients, controversial results are derived from different ABL kinase inhibitors, and more studies are needed to determine its therapeutic effect. Furthermore, the GECN-predicted targets, CSF2, IL-4, and IL-13, have been associated with COVID-19 in previous clinical studies [42-44]. GM-CSF is involved in the inflammatory phase, which induces a cytokine storm, and IL-4 and IL-13 are involved in the tissue repair phase, which causes pulmonary fibrosis. Therefore, clinical trials have focused on these three targets. Lenzilumab is a monoclonal antibody against GM-CSF with high binding affinity and low immunogenicity [45]. Lenzilumab neutralizes the GM-CSF effect and blocks signal transduction to myeloid progenitor cells. Lenzilumab has been shown to reduce the risk of death or respiratory failure in hospitalized COVID-19 patients [45]. Dupilumab is an IL-4Rα antagonist used in the treatment of atopic dermatitis, which blocks IL-13 and IL-4 signaling, may have potential as a treatment for COVID-19. A retrospective analysis of a trial of two-cohorts of patients infected with SARS-CoV2-confirmed the benefit with a lower risk of death in patients.

In addition, recent studies have developed innovative models for designing picomolar miniprotein inhibitors [4]. These models leverage computer-generated scaffolds designed to optimize target binding, folding, and stability around an ACE2 helix that interacts with the spike receptor binding domain or are docked against the RBD to identify new binding modes. These de novo design approaches offer a innovative and promising solution to the COVID crisis. However, clinicians have the critical timing of the pandemic outbreak, there is a dire need for potent therapeutics immediatly. Although designing drugs specific to SARS-CoV-2 is the gold standard solution, it requires complex and time-consuming procedures. Our GECN algorithm is built up with a knowledge database to further extend drug repurposing by summarizing information on FDA-approved drugs that could be applied immediately.

## 5. Conclusions

This study highlights the potential of using deep learning algorithms to discover potential therapeutic targets for COVID-19. Although our results were consistent with those of several studies, our predictions were difficult to confirm without more knowledge of virus-host interactions. The GECN identified several genes that may be targets for COVID-19 treatment, including ABL1, ABL2, CSF2, IL-4, and IL-13. Clinical trials have shown promising results for Lenzilumab, a monoclonal antibody against GM-CSF. However, more studies are needed to confirm the effectiveness of these drugs and their potential side effects. These findings provide a starting point for further research into potential treatments for COVID-19.

## Key Points

The graph-embedding convolution network (GECN), a novel deep learning model, was developed to identify COVID-19-related genes and druggable targets by analyzing COVID-19-related human protein interaction networks.

GECN identified several target genes, including ALB1/ALB2, CSF2, IL-4, and IL-13, that are meaningfully correlated with COVID-19, potentially involved in the entry of SARS-CoV-2 into host cells, cytokine storm, and pulmonary fibrosis.

The GECN algorithm can predict COVID-19 drug targets rapidly, accurately, and explainably, demonstrating its potential to expand our knowledge of disease pathologies and accelerate drug discovery at a low cost.

## Declarations

### Ethics approval and consent to participate

All authors had ethical approval and consent to participate.

## Consent for publication

All authors agreed with consent for publication.

## Availability of data and materials

The data that support the findings of this study are available from the corresponding author and listing authurs upon reasonable request.

## Author contributions

**Y-H.T.** conceptualized the study and wrote the methodology, original draft, and funding acquisition. **H.H.** wrote the original draft and reviewed and edited the manuscript. **C-N.Y.** contributed to the investigation, validation, formal analysis, and methodology. **S-H.W.** contributed to study investigation, validation, manuscript writing, review, and editing, and arranged resources. **O.K-S.L.** supervised the study, managed the project, and contributed to manuscript writing, review, and editing.

## Data Availability

The data supporting the findings of this study are available from the corresponding author upon reasonable request.

## Acknowledgements

The authors acknowledge financial support from the Ministry of Science and Technology. The Development and Construction Plan of the School of Medicine, National Yang-Ming University, now known as National Yang Ming Chiao Tung University, and Aiming for the Top University Plan, a grant from the Ministry of Education. We thank Muen Biomedical and Optoelectronics Technologies Inc. and all our colleagues at National Yang Ming Chiao Tung University for their encouragement and support.

---

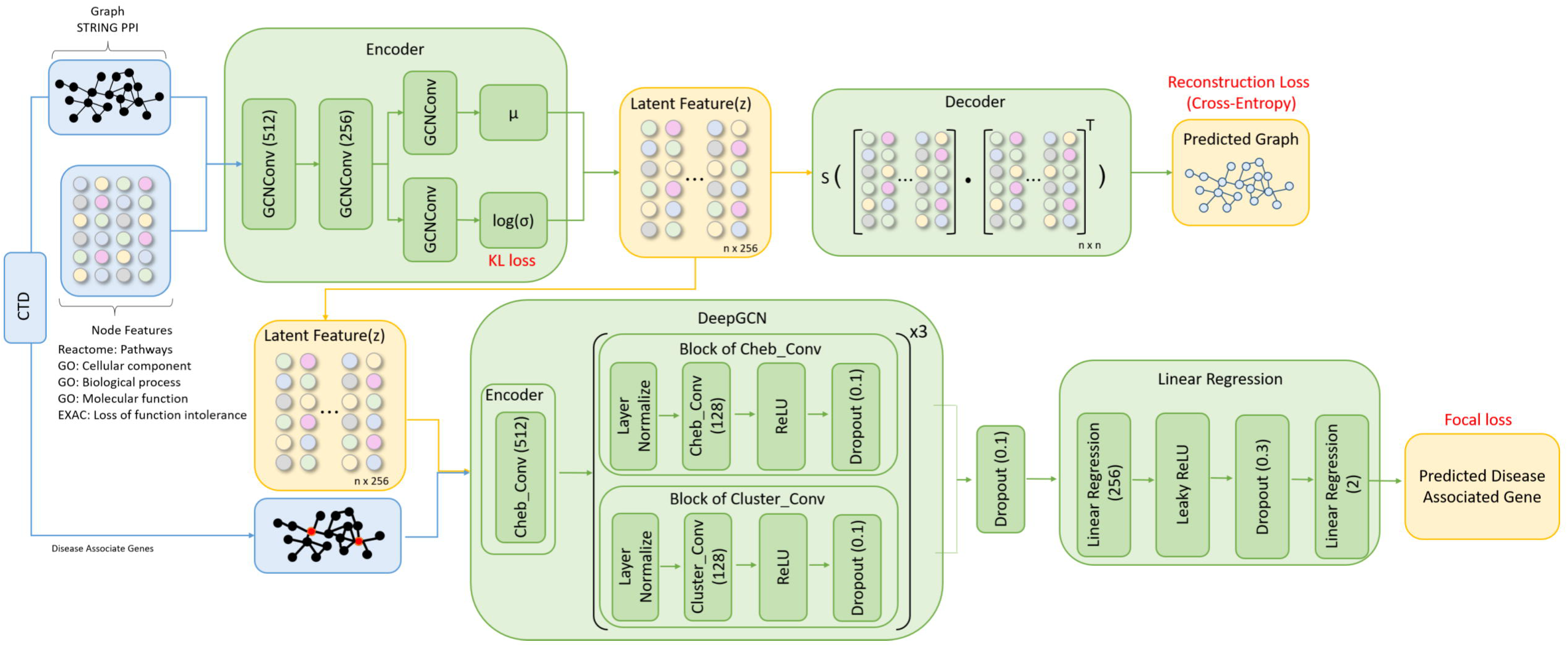

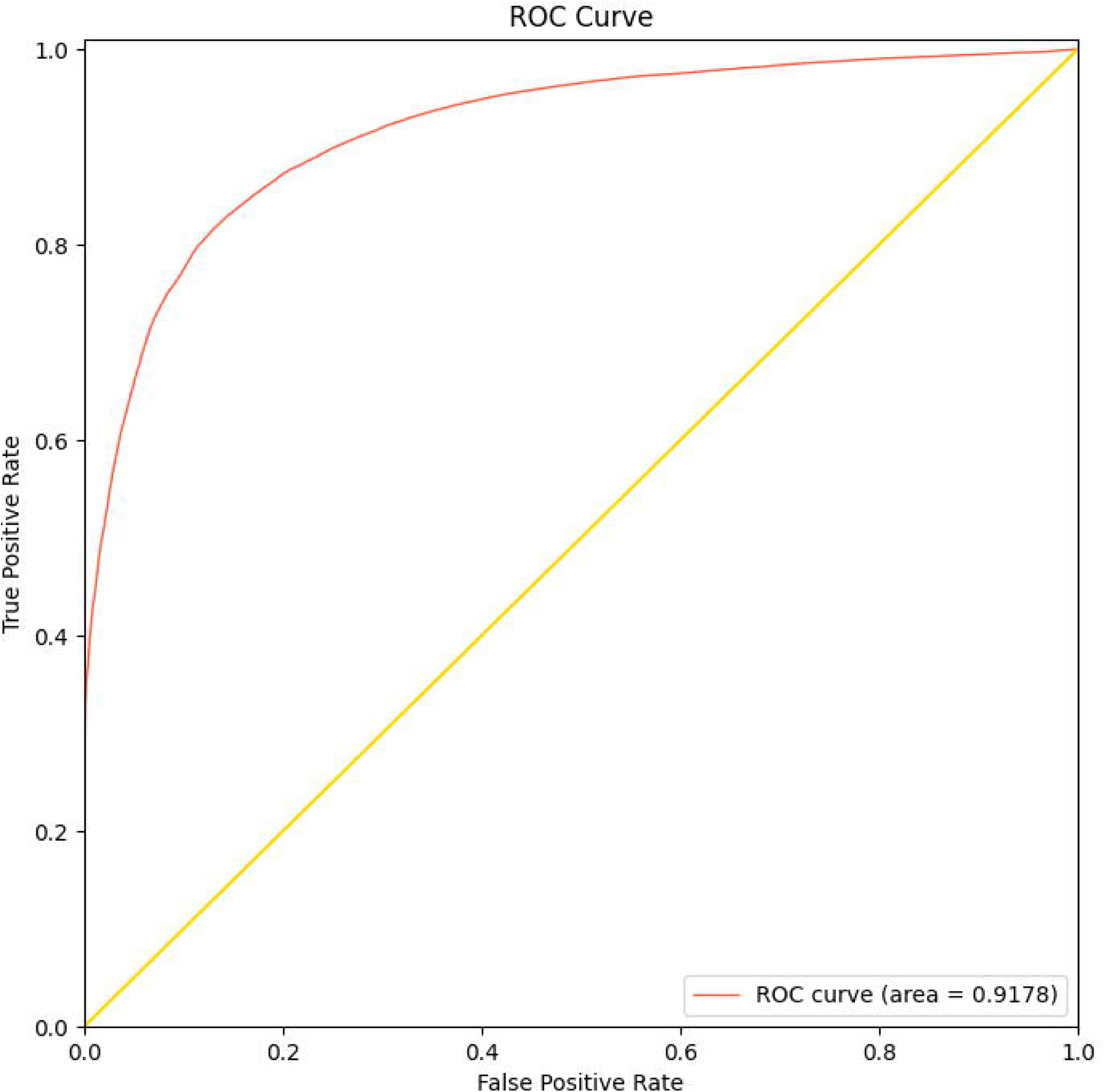

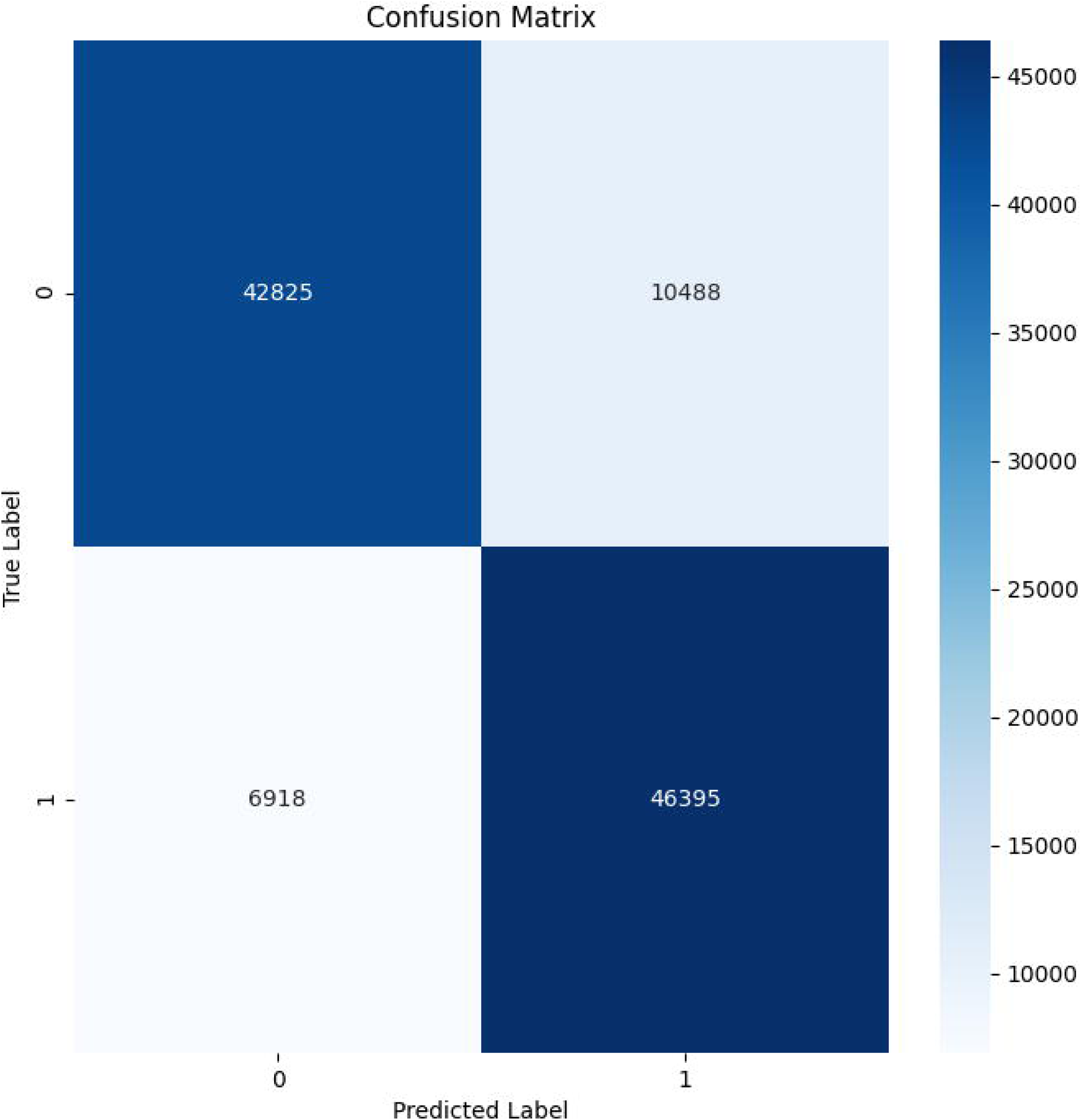

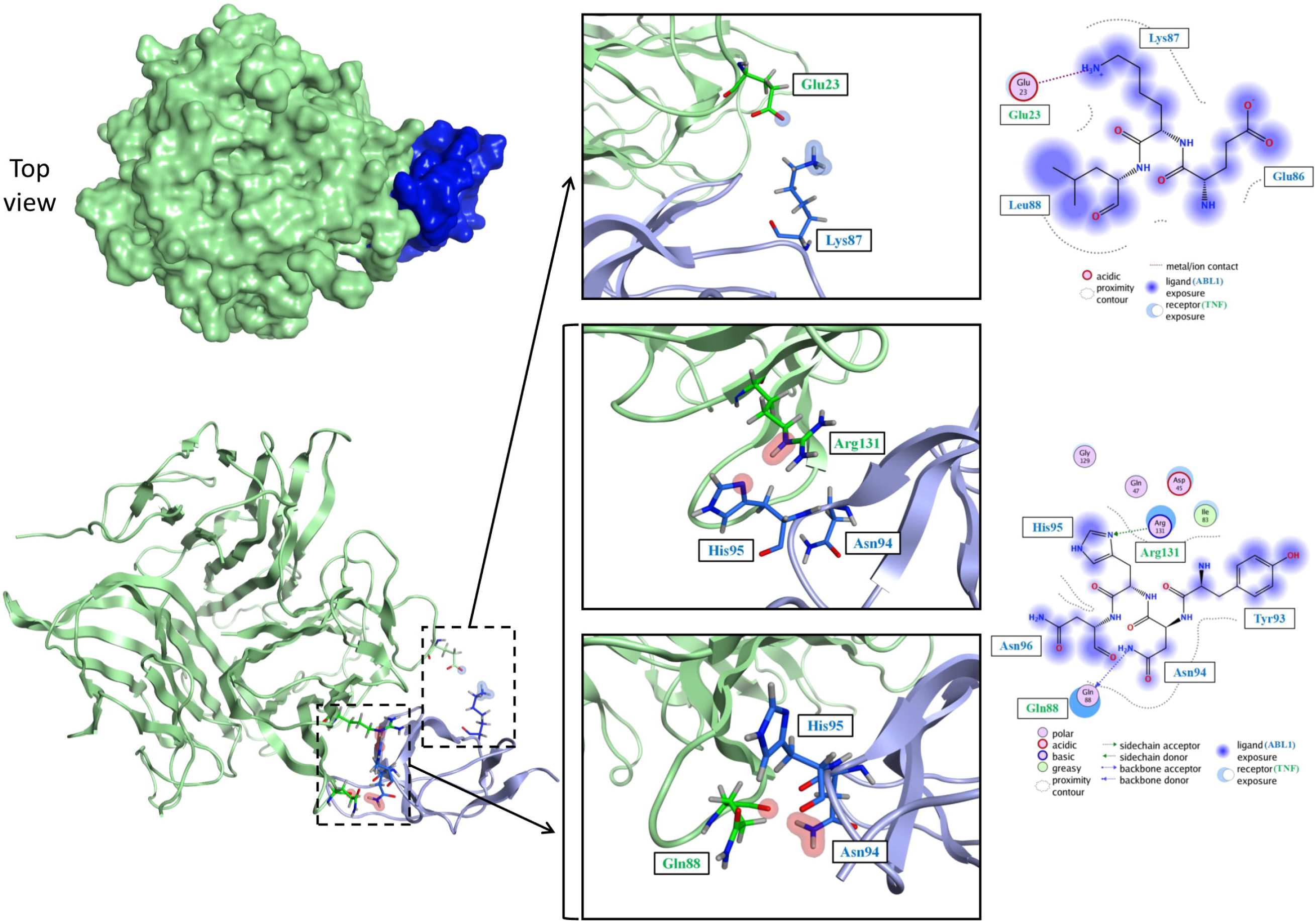

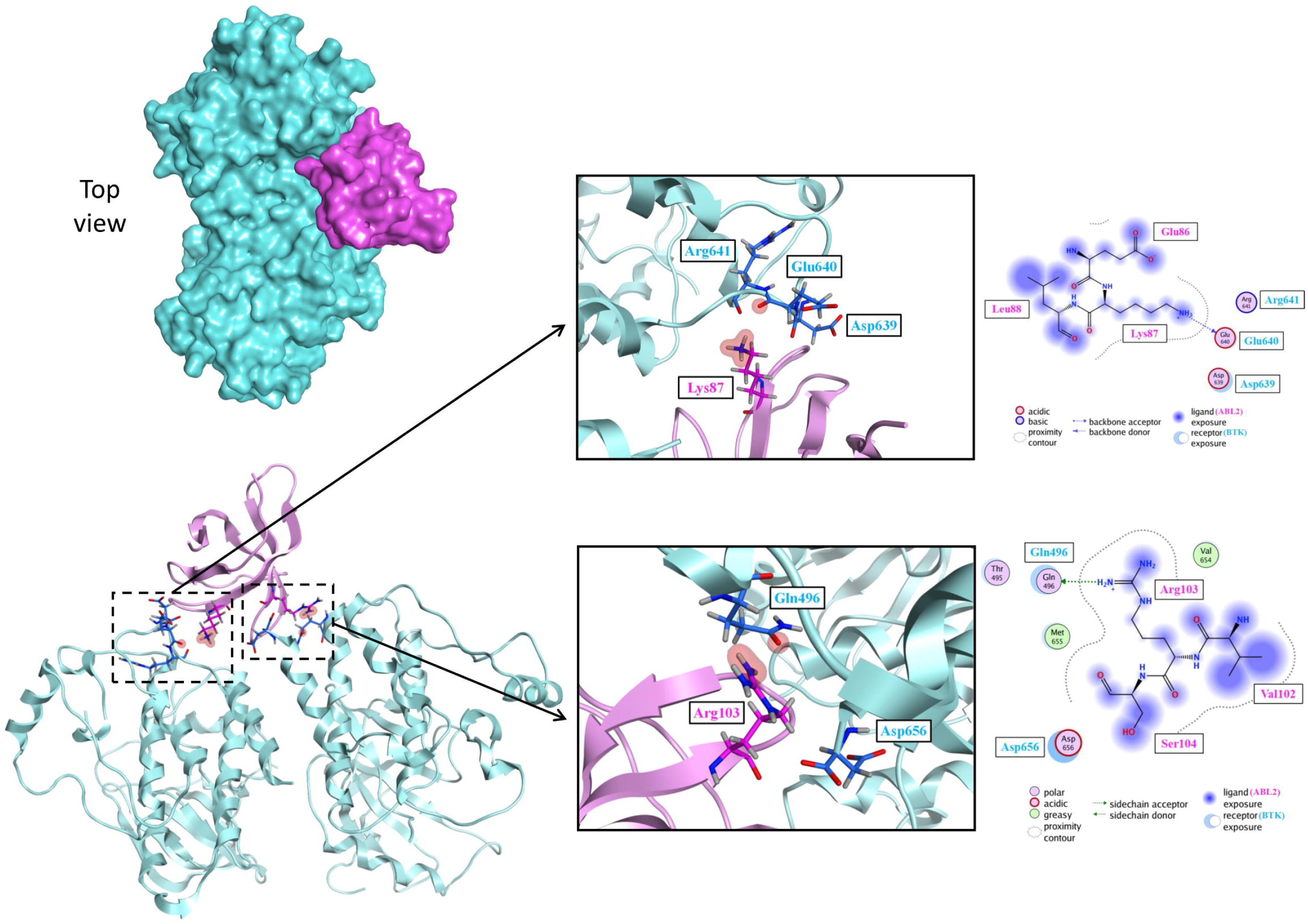

## References

1. Chen N, Zhou M, Dong X, Qu J, Gong F, Han Y, Qiu Y, Wang J, Liu Y, Wei Y et al: Epidemiological and clinical characteristics of 99 cases of 2019 novel coronavirus pneumonia in Wuhan, China: a descriptive study. Lancet 2020, 395(10223):507–513.

2. Ragab D, Salah Eldin H, Taeimah M, Khattab R, Salem R: The COVID-19 Cytokine Storm; What We Know So Far. Front Immunol 2020, 11:1446.

3. Singh TU, Parida S, Lingaraju MC, Kesavan M, Kumar D, Singh RK: Drug repurposing approach to fight COVID-19. Pharmacol Rep 2020, 72(6):1479–1508.

4. Cao L, Goreshnik I, Coventry B, Case JB, Miller L, Kozodoy L, Chen RE, Carter L, Walls AC, Park YJ et al: De novo design of picomolar SARS-CoV-2 miniprotein inhibitors. Science 2020, 370(6515):426–431.

5. Dauparas J, Anishchenko I, Bennett N, Bai H, Ragotte RJ, Milles LF, Wicky BIM, Courbet A, de Haas RJ, Bethel N et al: Robust deep learning-based protein sequence design using ProteinMPNN. Science 2022, 378(6615):49–56.

6. Scarselli F, Gori M, Tsoi AC, Hagenbuchner M, Monfardini G: The graph neural network model. IEEE transactions on neural networks 2008, 20(1):61–80.

7. Zhang XM, Liang L, Liu L, Tang MJ: Graph Neural Networks and Their Current Applications in Bioinformatics. Front Genet 2021, 12:690049.

8. Bruna J, Zaremba W, Szlam A, LeCun Y: Spectral networks and locally connected networks on graphs. arXiv preprint arXiv:13126203 2013.

9. Kipf TN, Welling M: Semi-supervised classification with graph convolutional networks. arXiv preprint arXiv:160902907 2016.

10. Veličković P, Cucurull G, Casanova A, Romero A, Lio P, Bengio Y: Graph attention networks. arXiv preprint arXiv:171010903 2017.

11. Yin W, Mao C, Luan X, Shen DD, Shen Q, Su H, Wang X, Zhou F, Zhao W, Gao M et al: Structural basis for inhibition of the RNA-dependent RNA polymerase from SARS-CoV-2 by remdesivir. Science 2020, 368(6498):1499–1504.

12. Beigel JH, Tomashek KM, Dodd LE, Mehta AK, Zingman BS, Kalil AC, Hohmann E, Chu HY, Luetkemeyer A, Kline S et al: Remdesivir for the Treatment of Covid-19-Final Report. N Engl J Med 2020, 383(19):1813–1826.

13. Ge Y, Tian T, Huang S, Wan F, Li J, Li S, Wang X, Yang H, Hong L, Wu N et al: An integrative drug repositioning framework discovered a potential therapeutic agent targeting COVID-19. Signal Transduct Target Ther 2021, 6(1):165.

14. Martin WR, Cheng F: Repurposing of FDA-Approved Toremifene to Treat COVID-19 by Blocking the Spike Glycoprotein and NSP14 of SARS-CoV-2. J Proteome Res 2020, 19(11):4670–4677.

15. Zeng X, Song X, Ma T, Pan X, Zhou Y, Hou Y, Zhang Z, Li K, Karypis G, Cheng F: Repurpose open data to discover therapeutics for COVID-19 using deep learning. Journal of proteome research 2020, 19(11):4624–4636.

16. Richardson P, Griffin I, Tucker C, Smith D, Oechsle O, Phelan A, Rawling M, Savory E, Stebbing J: Baricitinib as potential treatment for 2019-nCoV acute respiratory disease. Lancet 2020, 395(10223):e30–e31.

17. Zhou Y, Hou Y, Shen J, Kallianpur A, Zein J, Culver DA, Farha S, Comhair S, Fiocchi C, Gack MU et al: A Network Medicine Approach to Investigation and Population-based Validation of Disease Manifestations and Drug Repurposing for COVID-19. ChemRxiv 2020.

18. Horby P, Lim WS, Emberson JR, Mafham M, Bell JL, Linsell L, Staplin N, Brightling C, Ustianowski A, Elmahi E et al: Dexamethasone in Hospitalized Patients with Covid-19. N Engl J Med 2021, 384(8):693–704.

19. Fadaka AO, Sibuyi NRS, Madiehe AM, Meyer M: Computational insight of dexamethasone against potential targets of SARS-CoV-2. J Biomol Struct Dyn 2022, 40(2):875–885.

20. Goldman JD, Lye DCB, Hui DS, Marks KM, Bruno R, Montejano R, Spinner CD, Galli M, Ahn MY, Nahass RG et al: Remdesivir for 5 or 10 Days in Patients with Severe Covid-19. N Engl J Med 2020, 383(19):1827–1837.

21. Szklarczyk D, Gable AL, Lyon D, Junge A, Wyder S, Huerta-Cepas J, Simonovic M, Doncheva NT, Morris JH, Bork P et al: STRING v11: protein-protein association networks with increased coverage, supporting functional discovery in genome-wide experimental datasets. Nucleic Acids Res 2019, 47(D1):D607–d613.

22. Ashburner M, Ball CA, Blake JA, Botstein D, Butler H, Cherry JM, Davis AP, Dolinski K, Dwight SS, Eppig JT et al: Gene ontology: tool for the unification of biology. The Gene Ontology Consortium. Nat Genet 2000, 25(1):25–29.

23. Fabregat A, Sidiropoulos K, Viteri G, Marin-Garcia P, Ping P, Stein L, D’Eustachio P, Hermjakob H: Reactome diagram viewer: data structures and strategies to boost performance. Bioinformatics 2018, 34(7):1208–1214.

24. Alanazi A, Nojiri C, Noguchi T, Kido T, Komatsu Y, Hirakuri K, Funakubo A, Sakai K, Fukui Y: Improved Blood Compatibility of DLC Coated Polymeric Material. ASAIO Journal 2000, 46(4):440–443.

25. UniProt: a worldwide hub of protein knowledge. Nucleic Acids Res 2019, 47(D1):D506–d515.

26. Jassal B, Matthews L, Viteri G, Gong C, Lorente P, Fabregat A, Sidiropoulos K, Cook J, Gillespie M, Haw R et al: The reactome pathway knowledgebase. Nucleic Acids Res 2020, 48(D1):D498–d503.

27. Lek M, Karczewski KJ, Minikel EV, Samocha KE, Banks E, Fennell T, O’Donnell-Luria AH, Ware JS, Hill AJ, Cummings BB et al: Analysis of protein-coding genetic variation in 60,706 humans. Nature 2016, 536(7616):285–291.

28. Davis AP, Grondin CJ, Johnson RJ, Sciaky D, Wiegers J, Wiegers TC, Mattingly CJ: Comparative Toxicogenomics Database (CTD): update 2021. Nucleic Acids Research 2020, 49(D1):D1138–D1143.

29. Anno S, Hirakawa T, Sugita S, Yasumoto S: A graph convolutional network for predicting COVID-19 dynamics in 190 regions/countries. Front Public Health 2022, 10:911336.

29. Kingma DP, Ba J: Adam: A method for stochastic optimization. arXiv preprint arXiv:14126980 2014.

31. Wu Z, Pan S, Chen F, Long G, Zhang C, Philip SY: A comprehensive survey on graph neural networks. IEEE transactions on neural networks and learning systems 2020, 32(1):4–24.

32. Chiang W-L, Liu X, Si S, Li Y, Bengio S, Hsieh C-J: Cluster-gcn: An efficient algorithm for training deep and large graph convolutional networks. In: Proceedings of the 25th ACM SIGKDD international conference on knowledge discovery & data mining: 2019. 257–266.

33. Inc. CCG: Molecular operating environment (MOE). In.: Chemical Computing Group Inc. Montreal, QC, Canada; 2016.

34. Aleman MM, Walton BL, Byrnes JR, Wolberg AS: Fibrinogen and red blood cells in venous thrombosis. Thromb Res 2014, 133 Suppl 1(0 1):S38-40.

35. Clark AM, Labute P: 2D depiction of protein− ligand complexes. Journal of chemical information and modeling 2007, 47(5):1933–1944.

36. Sisk JM, Frieman MB, Machamer CE: Coronavirus S protein-induced fusion is blocked prior to hemifusion by Abl kinase inhibitors. The Journal of general virology 2018, 99(5):619.

37. Coleman CM, Sisk JM, Mingo RM, Nelson EA, White JM, Frieman MB: Abelson kinase inhibitors are potent inhibitors of severe acute respiratory syndrome coronavirus and Middle East respiratory syndrome coronavirus fusion. Journal of virology 2016, 90(19):8924–8933.

38. Dyall J, Coleman CM, Venkataraman T, Holbrook MR, Kindrachuk J, Johnson RF, Olinger Jr GG, Jahrling PB, Laidlaw M, Johansen LM: Repurposing of clinically developed drugs for treatment of Middle East respiratory syndrome coronavirus infection. Antimicrobial agents and chemotherapy 2014, 58(8):4885–4893.

39. Sauvat A, Ciccosanti F, Colavita F, Di Rienzo M, Castilletti C, Capobianchi MR, Kepp O, Zitvogel L, Fimia GM, Piacentini M: On-target versus off-target effects of drugs inhibiting the replication of SARS-CoV-2. Cell Death & Disease 2020, 11(8):656.

40. Cagno V, Magliocco G, Tapparel C, Daali Y: The tyrosine kinase inhibitor nilotinib inhibits SARS-CoV-2 in vitro. Basic & clinical pharmacology & toxicology 2021, 128(4):621–624.

41. Zhao H, Mendenhall M, Deininger MW: Imatinib is not a potent anti-SARS-CoV-2 drug. Leukemia 2020, 34(11):3085–3087.

42. Thwaites R: Sanchez Sevilla Uruchurtu A, Siggins MK, et al. Inflammatory profiles across the spectrum of disease reveal a distinct role for GM-CSF in severe COVID-19 Sci Immunol 2021, 6(57):57.

43. Donlan AN, Sutherland TE, Marie C, Preissner S, Bradley BT, Carpenter RM, Sturek JM, Ma JZ, Moreau GB, Donowitz JR: IL-13 is a driver of COVID-19 severity. JCI insight 2021, 6(15).

44. Shenoy S: SARS-CoV-2 (COVID-19), viral load and clinical outcomes; lessons learned one year into the pandemic: a systematic review. World Journal of Critical Care Medicine 2021, 10(4):132.

45. Temesgen Z, Assi M, Shweta F, Vergidis P, Rizza SA, Bauer PR, Pickering BW, Razonable RR, Libertin CR, Burger CD: GM-CSF neutralization with lenzilumab in severe COVID-19 pneumonia: a case-cohort study. In: Mayo Clinic Proceedings: 2020. Elsevier: 2382–2394.

