## Supplemental Table 1 for "Identification of targets for drug repurposing to treat COVID-19 using a Deep Learning Neural Network"

Supplementary Table 1

| Known COVID-19 related genes | New predicted interacted genes | Predicted probability |
|---:|---:|---:|
| TNF | ABL1 | 0.5188 |
| IL2 | ABL1 | 0.5166 |
| IL10 | ABL1 | 0.5145 |
| BTK | ABL2 | 0.5134 |
| IL1B | ABL1 | 0.5129 |
| IL2RA | ABL1 | 0.5123 |
| TNF | ABL2 | 0.5123 |
| IL2 | ABL2 | 0.5116 |
| IL7 | ABL1 | 0.5114 |
| BSG | ABCA2 | 0.5112 |
| TNF | A2M | 0.5112 |
| IL6 | ABL2 | 0.5107 |
| CSF3 | ABL1 | 0.5104 |
| IL2RA | ABL2 | 0.5099 |
| TNF | A1BG | 0.5098 |
| IL6 | AOC1 | 0.5098 |
| IL1B | A2M | 0.5098 |
| CXCL8 | ABL1 | 0.5096 |
| IL10 | ABL2 | 0.5096 |
| IL6 | ABCA1 | 0.5096 |
| CCL2 | ABL1 | 0.5095 |
| IL6 | ACAA1 | 0.5092 |
| IL1B | A1BG | 0.5089 |

**Table S1. New predicted links were further analyzed.** We filtered out new predicted links in the test set where one of the gene in a pair has known associations with COVID-19. The known associations are according to the “DirectEvidence” column in the CTD database.
